# A Systems-Based Framework for Immunisation System Design: Six Loops, Three Flows, Two Paradigms

**DOI:** 10.1101/2021.07.19.21260775

**Authors:** Catherine Decouttere, Nico Vandaele, Kim De Boeck, Stany Banzimana

## Abstract

Despite massive progress in vaccine coverage globally, the region of sub-Saharan Africa is lagging behind and not on track for Sustainable Development Goal 3 by 2030. Sub-national under-immunisation, related to geographical and social heterogeneity, is part of the problem. System-wide changes could transform current immunisation systems to become more sustainable, resilient and inclusive. A framework is proposed that captures the complexity of immunisation systems and offers directions for sustainable redesign. Insights were extracted from literature, stakeholder workshops, and field research in Rwanda and Kenya. A conceptual model of the national immunisation system was co-developed and validated with stakeholders. Leverage points were suggested for intervention scenario building. The Immunisation System Diagram assembles the paradigms of planned and emergency immunisation in one system and emphasizes the synchronized flows of vaccine receiver, vaccinator and vaccine. Six feedback loops capture the main subsystems. Sustainability and resilience are assessed based on loop dominance and dependency on exogenous factors such as donor funding and vaccine stockpiles. In group model building workshops, the diagram invites stakeholders to share their mental models, to assess the system’s performance and to trigger detection of root causes and leverage points. The framework provides a systems-approach for problem structuring and policy design.

## Introduction

Since the introduction of routine childhood immunisation in low-income-countries (LICs) in 1974 as the Expanded Program on Immunisation (EPI), a spectacular global decrease in neonatal mortality rate (NMR) and under-five mortality rate (U5MR) has been observed. However, despite massive efforts in EPI, the world-wide diphtheria-tetanus-pertussis third dose (DTP3) national immunisation coverage level has nearly stagnated since 2009 at 85%. In 2018, the DTP3 coverage in the African continent was only 76% (United Nations Inter-agency Group for Child Mortality Estimation, 2018; WHO-Unicef, 2019), which is well below the 2015 target of the Global Vaccine Action Plan (GVAP) of 90% as shown in Figure 1.

**Figure 1.**
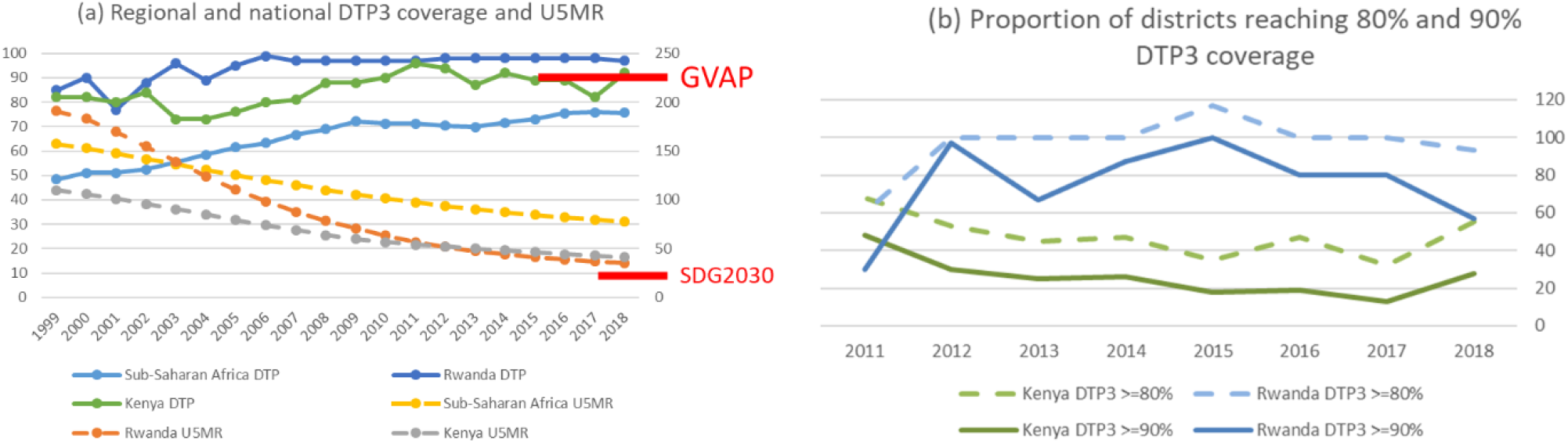
SSA and national coverage level of 3rd dose DTP vaccine and Under-five mortality rate. GVAP 2015 national target of 90% was feasible for Rwanda and Kenya but not for most SSA countries. (b) Even in countries where the national target was met, there is great subnational diversity: only 28% (Kenya) and 57% (Rwanda) of the districts reached 90% coverage levels for DTP3 that lead to herd immunity. Data from WHO/UNICEF Joint Reporting Form. (www.who.int/immunization/monitoring_surveillance/data/en/)

At the subnational level, the 2020 GVAP target of 80% DTP3 coverage in every district seems unlikely for most sub-Saharan (SSA) countries. Although high-quality sub-national indicators are not always readily available for SSA, estimates of U5MR based on different sources show significant variations and point out that local conditions play a more critical role than administrative boundaries and that major health inequities exist which penalize the poor people in rural areas and urban slums (Ndwandwe et al., 2018). According to a Sustainable Development Goal (SDG) baseline analysis for NMR and U5MR in Africa, many localities and geographical regions will need specifically targeted interventions if they are to reach the health-related SDG targets (Golding et al., 2017).

Furthermore, a Grand Convergence is expected, in which an increasingly pressing *Double burden of disease* is observed in SSA. Non-communicable chronic conditions, related to lifestyle, climate change, and demographic shift, are catching up with infectious diseases and malnutrition as causes of morbidity and mortality (Levine, 2017).

According to the Lancet Commission on the future of health in SSA (Agyepong et al., 2017), a framework shift is required in the pursuit of sustainable health outcomes and new health systems should focus on local context, people-centredness and prevention. Similarly, both Immunization Agenda 2030 (IA2030) and the GAVI 5.0 plan urge for both inclusive systems, viewing immunization from a life-course perspective (GAVI The Vaccine Alliance, 2019; WHO, 2019) and for resilient systems which are resistant to both chronic stress and acute shocks, and that show the ability to adapt and transform into more resilient systems (Barasa, Mbau, & Gilson, 2018).

Some subsystems of the immunisation system (IMS), such as the vaccine supply chain, have been intensively modelled and a number of studies traced main challenges and their root causes to identify key areas of intervention in the particular subsystems (De Boeck, Decouttere, & Vandaele, 2019; Duijzer, Jaarsveld, & Dekker, 2018; Haidari et al., 2017; Lemmens, Decouttere, Vandaele, & Bernuzzi, 2016; Wedlock et al., 2018; Yadav, 2015). In addition, the impact of the vaccine supply chain performance, measured by vaccine availability, on reported IMS performance, measured by immunisation coverage, was modelled (Gooding, Spiliotopoulou, & Yadav, 2019). On a separate line, human behavioural impact such as the role of vaccine hesitancy has been studied (Larson et al., 2018). Systems thinking and implementation research, applied to Low- and Middle-Income Countries’ (LMICs) health systems, highlighted the power of taking a system’s perspective to understand complexity, adaptive behaviour and root causes of missed opportunities to vaccinate (Adamu et al., 2019; Ozawa, Paina, & Qiu, 2016; Remme et al., 2010; Semwanga, Nakubulwa, & Adam, 2016; “WHO | The Alliance for Health Policy and Systems Research (AHPSR),” 2019). Dynamic simulation models and health economic decision frameworks further stress the importance of transdisciplinary phenomena, capturing delays and non-linear behaviour for decision making (Auping, Pruyt, & Kwakkel, 2017; Baltussen et al., 2017; Gonçalves, 2011; Lee, Mueller, & Tilchin, 2017; Rouwette, Vennix, & Felling, 2009).

However, the combined, dynamic and long-term effect on health outcomes and SDGs has not directly been addressed with the objective of IMS redesign. Furthermore, considering implementation aspects when designing interventions for sustainable and resilient immunisation systems, has largely been unaddressed.

The aim of this paper is (1) to develop a systems-inspired framework, capable of linking system actors and interventions to the health-related SDGs and (2) to support stakeholders in defining design directions and implementation trajectories for a resilient and sustainable LMIC IMS.

## Materials and methods

Because the targeted framework is intended for the redesign of a complex health system, suitable system design processes were first screened. To this end, peer-reviewed papers discussing LMIC IMS design, vaccine supply chain design, and health systems strengthening were analysed. Additionally, grey literature from global immunisation agencies, including WHO, UNICEF and GAVI were reviewed.

Based on their immunisation performance, their current and future challenges, and data availability within an existing collaboration opportunity, the immunisation systems of Rwanda and Kenya were selected to provide the context for the development of the framework. Background data were gathered from policy documents such as the “comprehensive Multi-Year plan for Immunization” (cMYPs).

The framework design process (Figure 2), consists of iterative cycles of stakeholder engagement in the stages of exploration, conceptualisation, testing and validation which took place between 2016 and 2019. An overview of the stakeholder engagement activities and methodologies applied is provided in in Table 1.

**Table 1.** Overview of stakeholder interaction activities that served as input for IMS diagram development

| Interaction type |  | Qualitative research method | Participants, location, timing | Main topics discussed |
| --- | --- | --- | --- | --- |
| 1 | Exploration **global** level | Stakeholder interviews, conference workshops | 1a MSF, Brussels<br>1b UNICEF, SC division, Copenhagen<br>1c Vaccine manufacturers<br>1d Donor organisations<br>2015- 2019 | Links between Health system development and humanitarian aid, infrastructure gap, skills retention, project handover<br>Vaccine funding, humanitarian mechanisms, vaccine stockpiles<br>Health systems strengthening approach UNICEF, GAVI<br>Improvement potential at interfaces of disciplines, e.g. vaccine manufacturing & procurement conditions, temperature requirements |
| 2 | Mental model elicitation **national** immunisation system | Group model building | Senior and middle EPI staff national level. Mixed groups:<br>Kigali 2016:<br>22 participants from Rwanda, Kenya, Uganda, Tanzania, South-Sudan<br>Nairobi 2017:<br>25 participants from Rwanda, Kenya, Uganda, Burundi, Tanzania, South-Sudan | Stakeholders involved in execution and decision making<br>Mental model change from Supply Chain to systems thinking<br>Policy making in cMYP, immunisation plan<br>Private sector role and partnerships (logistics)<br>Sharing views between countries, adapt approach to country<br>Financial mechanisms, equity, sustainability, efficiency<br>Data management and issues, denominator error<br>Vaccine coverage and health outcomes: delays, epidemiology<br>Implementation complexity: new vaccines, eLMIS<br>Workforce capacity, immunisation integration in health services |
| 3 | Mental model elicitation **district** level | Interviews, observation, numerical data collection at DHs | Rwanda: 2 district hospitals (2018-2019)<br>Kenya: 2 district hospitals (2018) | Vaccine supply chain, vaccine wastage, ordering process<br>Outreach planning and funding<br>Skills and training EPI staff and Community Health Workers<br>Diversity between HCs in the district, root causes of coverage variation<br>Demand data availability, district coordinating role |
| 4 | Mental model elicitation **local** level | Interviews, observations, numerical data collection at HCs, outreach, CHWs | <u>Kenya</u>: 5 health facilities (2018)<br><u>Rwanda</u>: 5 health facilities (2019) | Local vaccine management organization, vaccine transport<br>Community health worker, defaulter tracing, disease case detection<br>Demand-side determinants, access to HFs<br>Impact of local problem-solving capacity at HF |
| 5 | Validation **national** level CLD | Feedback workshops | EPI, WHO, implementers<br>Nairobi 2018, Kigali 2019 | Data management<br>Public-Private health services |
| 6 | Validation **national** level CLD | Feedback meetings | EPI, Kigali, 2019<br>UNICEF, Kigali, 2019 | Financial sustainability<br>Supply chain redesign, Digital transformation<br>Disease surveillance, emergency immunisation (measles, Ebola)<br>Integration with other preventive HC systems (nutrition, Primary HC) |
| 7 | Validation and refinement by international domain experts | Feedback meetings (2019) | 7a Academic anthropologist<br>7b Funding mechanism expert<br>7c Academic Public Health<br>7d Academic systems expert | Vaccine hesitancy, emergency immunisation<br>Design thinking, paradigm shift, Innovation interventions<br>Sustainable mechanisms, Local contextual approach, community health<br>Sustainability and resilient systems, systems levels and interventions |

**Figure 2.**
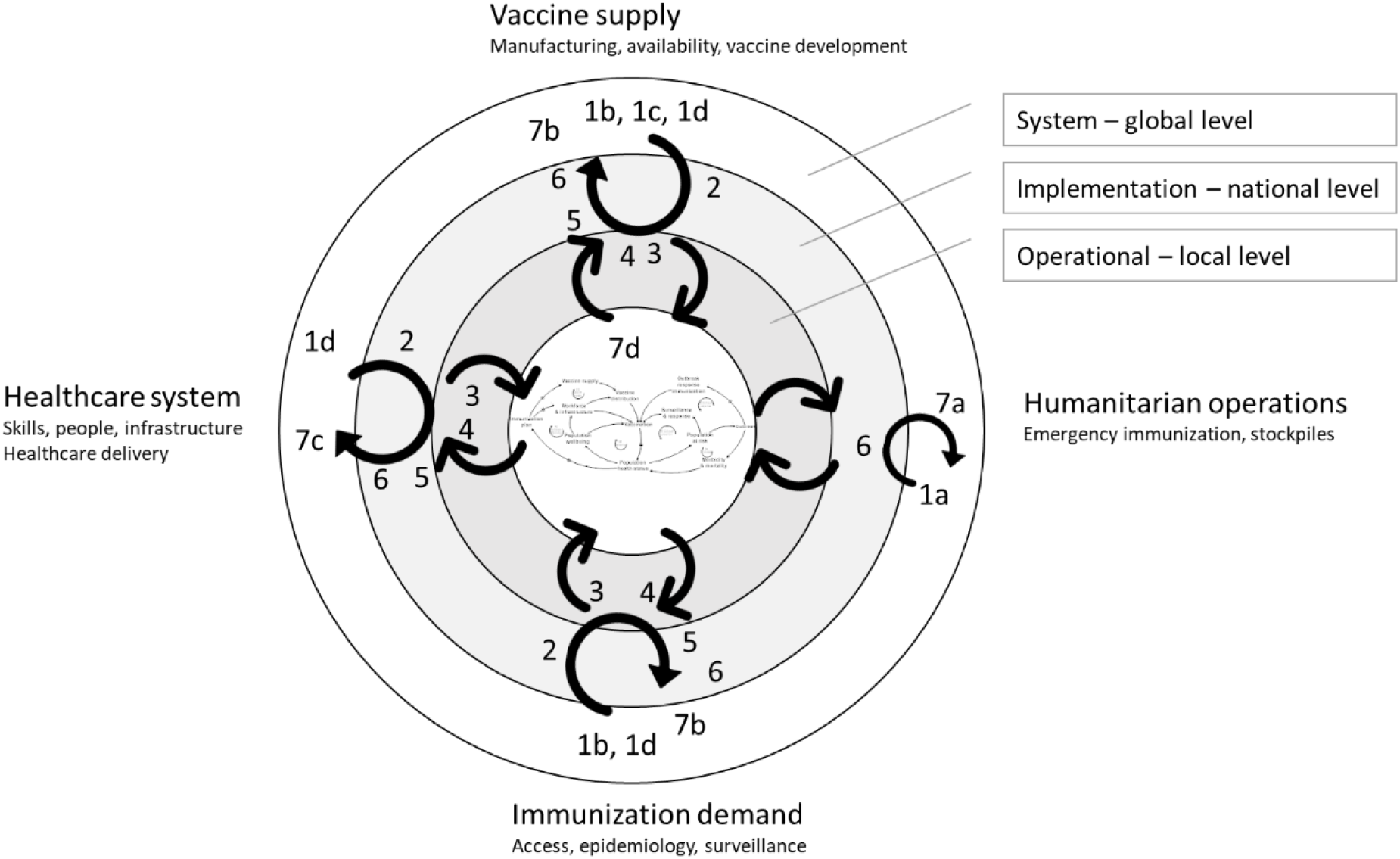
Map of interaction activities with stakeholders for the development and validation of the systems diagram. The numbers refer to the stakeholder engagement activities listed in Table 1. The concentric rings, with the resulting IMS diagram in the centre, indicate the system levels at which the stakeholder operates, or has his or her expertise. The four outer orientations (Vaccine supply, Humanitarian operations, Immunisation demand and Healthcare system) refer to the main background of the stakeholders. One activity can appear at different locations, for example a group model building session with stakeholders with different backgrounds. The grey arrows show the iterative, multi-level knowledge build-up while the black arrows indicate how this knowledge flows to and from the IMS diagram under construction

Applying principles from systems thinking and complexity science, it was demonstrated how delays, feedback loops, and key leverage points are visually represented in a highly aggregated causal-loop-diagram (CLD). Such a diagram has been shown to align and refine the mental models of stakeholders to enable them to identify root causes and potential avenues for system redesign (Kopainsky, Hager, Herrera, & Nyanga, 2017; Sterman, 2000). System interventions can be most effective when they are acting on system leverage points (Meadows & Wright, 2008). The identification of these leverage points and the orientation in which to direct them, is facilitated by combining the expertise of stakeholders and systems modellers. In addition to the IMS diagram, a framework for intervention scenario building was generated, based on the 5-level “Intervention Level Framework” (Malhi et al., 2009).

## Results

### Framework requirements

Aiming at broad applicability, the framework should fit into a broader system design or systems modelling process, such as the five-step framework for system design, the iterative system dynamics modelling process, a community-based system dynamics project, or a human-centred design process (Decouttere, Vandaele, Lemmens, & Bernuzzi, 2016; Hovmand, 2014; Sterman, 2000).

First, the framework must invite stakeholders to **share their mental models with other stakeholders**. The framework reveals the structures that drive the system’s dynamic behaviour: feedback loops, delays, and accumulation. For example, the framework should help an EPI vaccine procurement officer at the national level, an EPI nurse in a health centre (HC) and a community leader to understand how they are connected within the system by the vaccines propagating through the supply chain, the number of people living in the community and the vaccination coverage targets. In addition, it should explain to them how vaccine-preventable disease (VPD) outbreaks occur and each stakeholder’s role in preventing outbreaks

Second, the framework must enable stakeholders to **identify root causes** and to **co-design interventions** around **key leverage points**. Building further on the previous example, the framework should reveal to the procurement officer, the nurse, and the community leader some of the root causes of previously observed missed-opportunities-to-vaccinate (MOVs), such as a vaccine out-of-stock, and its root cause. It should also empower stakeholders in contributing to the design of sustainable solutions and avoiding the introduction of another problem into the system.

The framework is built around causal loop diagrams (CLDs) which show causal relationships between system actors and highlight reinforcing (R) or balancing (B) feedback loops. CLDs show the direction of the causality (+/- sign) and indicate delays.

### IMS: two paradigms, three flows, six loops

The goal of the IMS is captured by SDG3: “Ensure healthy lives and promote well-being for all at all ages “(UN, n.d.). More specifically, the IMS should deliver the SDG3.2 targets on NMR (<12/1000 live births) and U5MR (<25/1000 live births) by 2030. In line with SDG3, global and national intermediate targets have been set, and programs for immunisation have been defined: the Global Vaccine Action plan (GVAP), Decade of Vaccines (DoV), disease-specific eradication and elimination programs, and Immunization Agenda 2030. Central to the IMS is the strategy of **vaccination** to ensure health and well-being. The combined effect of vaccination and complementary health-promoting strategies, such as nutrition, is observed in the reduction of U5MR.

#### Two paradigms meet: planned and emergency immunisation

Following the purpose of the IMS, the implementation is materialized in the National Immunisation Program (NIP) which is based on three pillars: Routine Immunisation (RI), Supplemental Immunisation Activities (SIAs) and surveillance for epidemic-prone diseases. Both RI and SIAs are intended to pro-actively prevent VPD cases. However, when disease cases occur, they are detected by the surveillance mechanism which activates a response mechanism in order to prevent a wider outbreak. If, despite the activation of this mechanism, the outbreak reaches a certain scale and the local or national IMS is no longer able to control the health risk, outbreak response immunisation (ORI) can be invoked in order to comply with International Health Regulations (IHR) to guarantee international health safety (World Health Organization (WHO), 2016; World Health Organization (WHO) & Centers for Disease Control and Prevention (CDC), 2010). In the aggregated system diagram (Figure 3) Planned and Emergency Immunisation are represented as two complementary pathways in the system, that are connected by *Vaccination*, the common strategy, and *Population Health Status*, the common goal.

**Figure 3.**
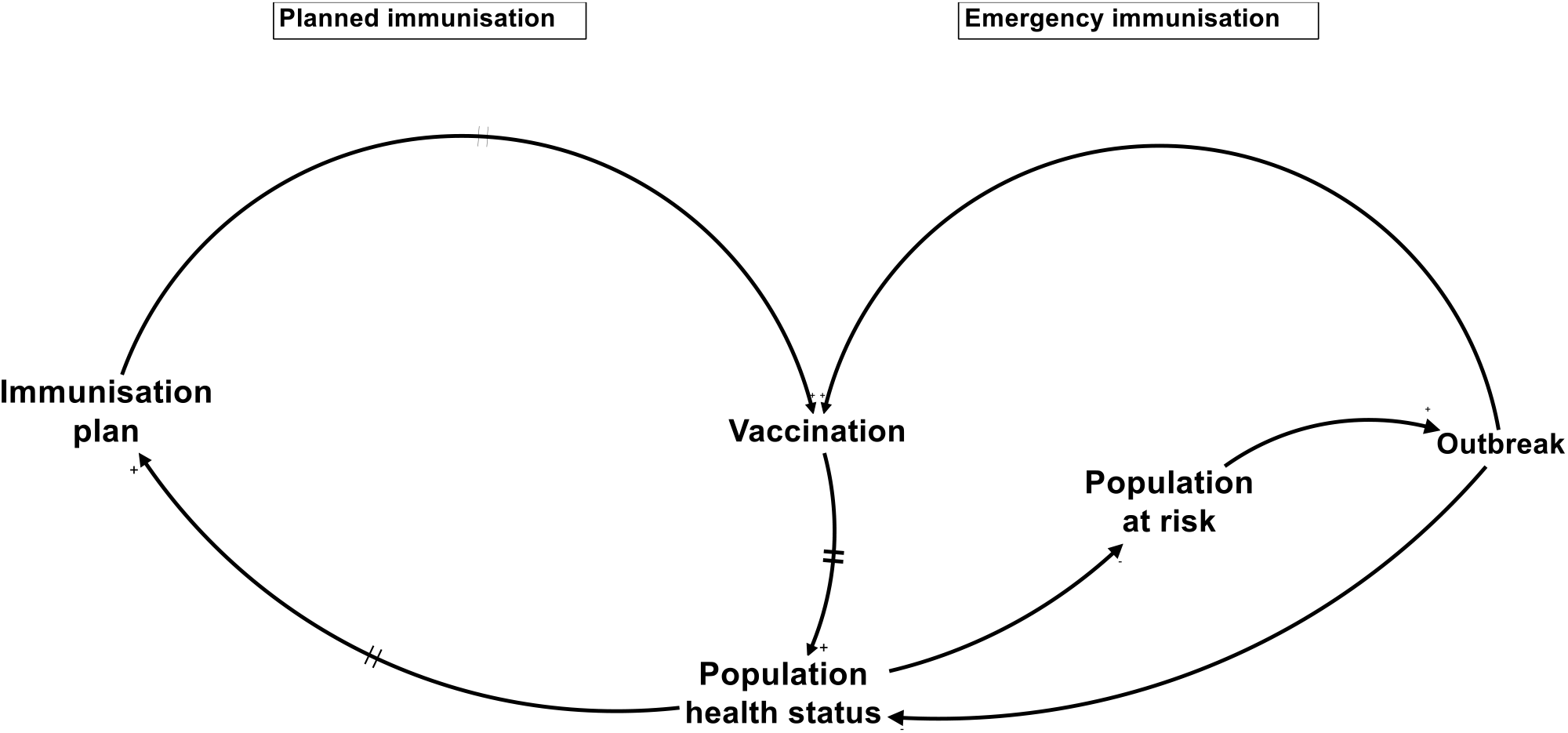
Aggregated system diagram highlighting two paradigms: planned and emergency immunisation.

At the national level, *Planned immunisation* is guided annually by the NIP that defines yearly domestic and donor budgets. RI involves continuous vaccine services roughly proportional at the birth rate, while SIAs, or preventive campaigns, aim to reach a large number of people from a target population in a short period of time. SIAs are planned according to disease-specific programs, including eradication programs (Global Polio Eradication Initiative (GPEI), Measles & Rubella Initiative (MRI) following specific guidelines (WHO Regional office Africa, 2015)). Another campaign-like format is Periodic Intensification of Routine Immunisation (PIRI) (World Health Organization, 2016), aimed at rapidly increasing coverage levels in communities that are insufficiently reached by RI. The dynamics between RI and SIAs differ significantly: RI is a generic, efficiency oriented immunisation strategy that leads to a stable population protection, whereas SIAs aim to quickly elevate immunisation levels. Both RI and SIAs are included in the NIP and require the supply, distribution and administration of vaccines which are largely performed partly by the same actors and partly by dedicated actors, both included in the *Planned immunisation*.

*Emergency immunisation* is driven by a different paradigm: reaction to minimize damage. Actors, resources, and policies differ as well, since international emergency response is guided by IHR and WHO-led emergency decision frameworks (World Health Organization, 2017b) for vaccination in acute humanitarian emergencies. Equally as in the case of domestic emergency response, the approach is campaign-based and much more time-limited and case- and disease-specific, and it has a higher degree of uncertainty, such as the timely securing of vaccines from emergency stockpiles.

Although *Planned* and *Emergency Immunisation* act as complementary mechanisms, *Planned Immunisation* is preferred since it efficiently prevents VPD cases and strengthens the population health status in accordance with the SDGs. When it comes to priority setting and funding allocation, however, it is shown that treatment of patients typically prevails over prevention (Bishai, Paina, Li, Peters, & Hyder, 2014; Tversky & Kahneman, 1974). To counteract this, The WHO/UNICEF-EPI program fosters the GVAP and the DoV acceleration plans with adequate funding from GAVI and other donors. These plans have successfully separated decisions on national *Immunisation plans* from other health care decisions and enabled LMIC governments to commit to immunisation. The required vaccination coverage targets for most pathogens lead to herd immunity. When the coverage level drops below this threshold it leads to outbreaks and activates the need for emergency immunisation.

The risk of a VPD outbreak depends on the combined epidemiological risk factors having an effect at **the community level**. Vaccination coverage, living conditions, exposure to pathogens and previous infections at the local level amongst others can deviate dramatically from the district or national average levels.

*Planned* and *Emergency immunisation* are usually designed and modelled in a different context: the former in the framework of health systems strengthening and disease-specific programs (including eradication), and the latter in outbreak response (including humanitarian operations). However, a systems perspective inevitably leads to their connection points where they impact each other in their need for local resources and their effect on *Vaccination* and *Population Health*. To redesign towards more efficient, resilient and sustainable IMSs, both approaches need to be considered as parts of one IMS.

#### Three flows - vaccinee, vaccinator and vaccine - meet each other across interacting feedback loops

*Vaccination* is the central point where a healthy **person** receives a potent **vaccine** administered by a skilled **health professional**. These three flows need to converge at the right moment in order to generate the desired immune response, both in planned and emergency immunisation. The location may be fixed (health centre, HC), outreach or mobile. To this end, vaccines are carefully transported to the vaccination point, nurses are trained and need to be available, vaccination sessions are organized and the mothers or targeted populations attend the sessions, sometimes invited by community health workers.

#### Six loops

The main mechanisms that drive the IMS are captured by six feedback loops, as indicated in the IMS CLD (Figure 4). On the *Planned immunisation* part of the system, *Vaccination* is at the intersection of three reinforcing feedback loops, each of them corresponding to 3 flows: (R1) the *Population* w*ellbeing* loop (child), (R2) the *Workforce and Infrastructure* loop (nurse), and (R3) the V*accine supply* loop (vaccine). At the *Emergency immunisation* side of the system, two balancing feedback loops represent the immunisation of people at risk before a wider outbreak occurs (*Surveillance and response loop*, B1), or during an outbreak which could not be controlled by the national IMS (*Outbreak response immunisation loop, B2*). Finally, a reinforcing *Outbreak* loop shows disease spreading among people at risk during an outbreak (R4). Evidently, this diagram is layered with respect to the individual pathogens and are connected at several shared system elements. A detailed explanation of the loops can be found in Annex 1.

**Figure 4.**
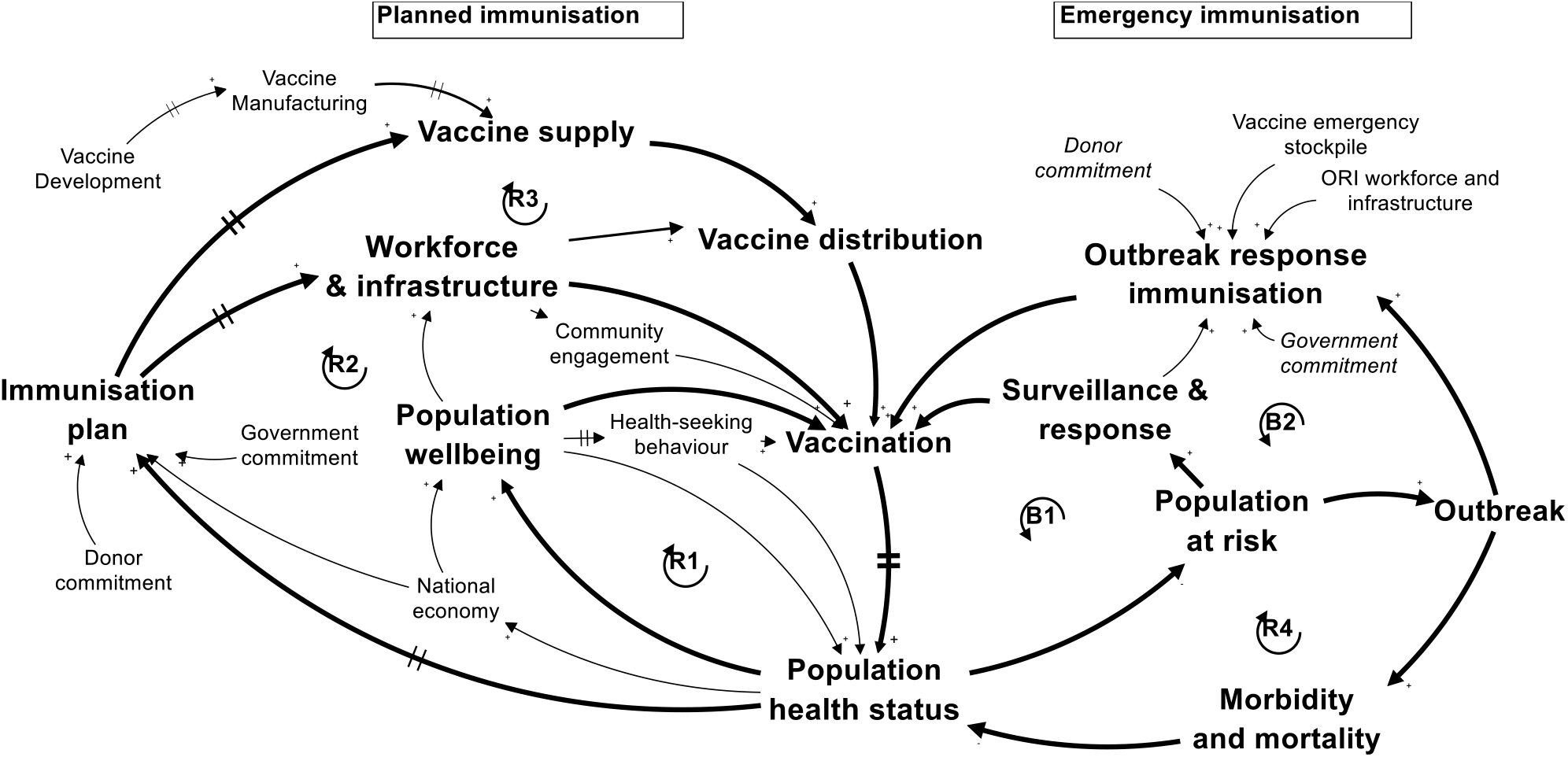
IMS diagram: Aggregated causal-loop diagram of IMS in a low- and middle-income country. Applicable to national, subnational level and local settings.

### A tool for system redesign

The application of the systems approach framework based on the IMS diagram is illustrated as a 3-phazed iterative process: (1) creation of a shared insight on how the IMS works, (2) evaluation of current system performance, sustainability and resilience, and (3) directions for sustainable and resilient system redesign.

#### Shared system insight

The redesign process first involves stakeholder **identification** (Decouttere et al., 2016). The IMS diagram facilitates this by providing a generic framework for stakeholder selection to engage in group model workshops and co-creation sessions (Figure 5).

**Figure 5.**
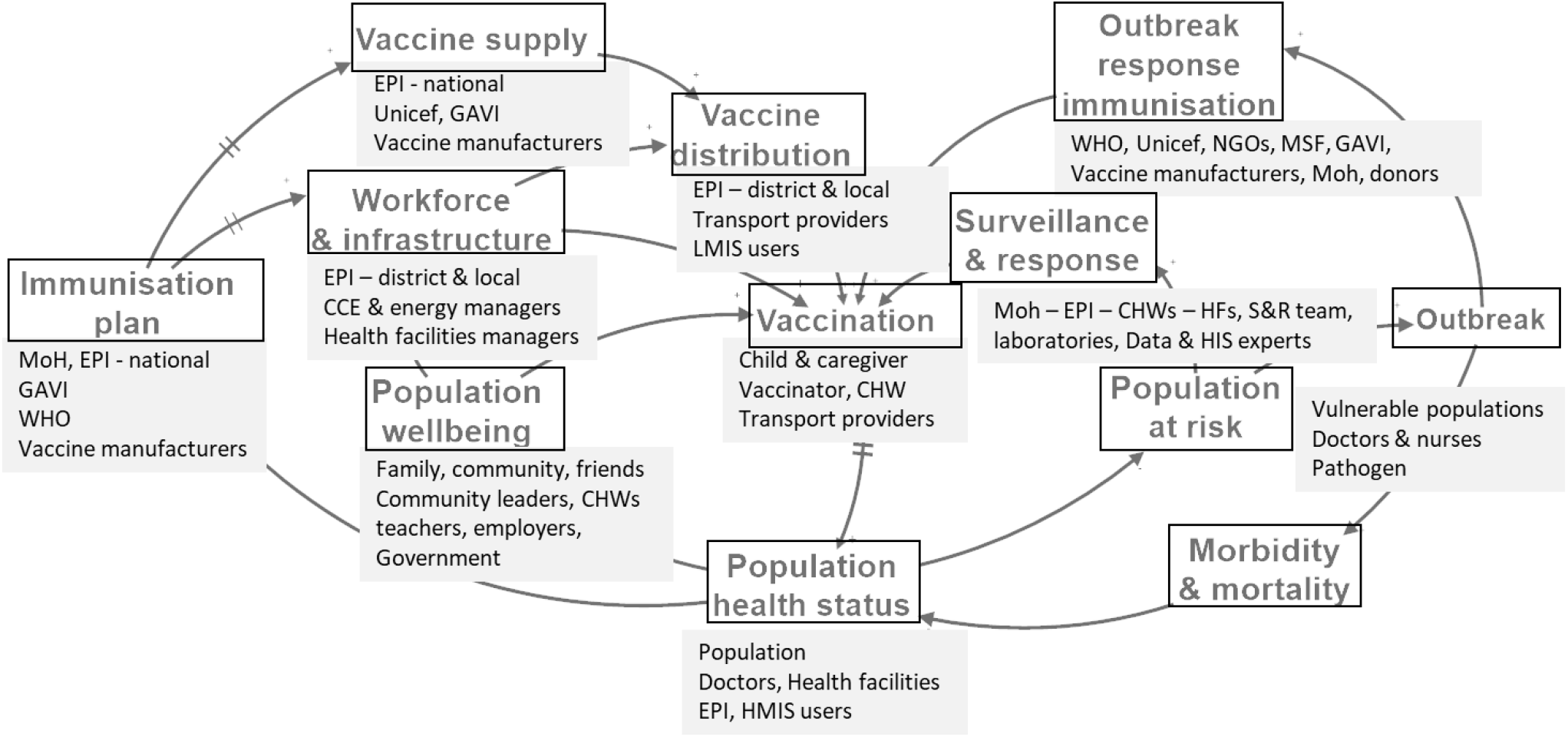
Stakeholders mapped on the generic LMIC IMS diagram.

Each system element in the IMS diagram represents specific stakeholders. By inviting stakeholder subsets from different levels in Rwanda and Kenya to collectively clarify the system’s working and performance, **shared insights** arose and the stakeholders’ mental models were broadened and refined, similar to the findings of Kopainsky et al. (2017). Firstly, a system level insight gained was the enhanced awareness of the child, nurse and vaccine flow synchronization, and its impact for planning and evaluating the interventions, including demand generation. Secondly, the long delays, causing rigidity and potential instability in the vaccine supply chain, were pointed out. Thirdly, the role of human decision making and the adaptive behaviour of the system was regarded to broadly impact the performance of the system, and seemed to culminate at the point of vaccination.

#### Current system assessment

Starting from behaviour-over-time graphs related to *Vaccination* and *Population Health Status such as Figure 1*, the stakeholders in Rwanda and Kenya were triggered to recall their experiences and share their perspectives on the factors driving the current system. A structured discussion supported by the IMS diagram yielded the following three main insights. First, the different contextual factors at **subnational level** and the resulting disparities in local vaccination coverage and health outcomes confirmed the need to refine system design below the national level. Using the IMS diagram as a template for mapping the different local contexts with their versions of dominating feedback loops, proved to be a powerful communication tool that drew attention to the contrasting elements. A typical example is the different accessibility issues in a remote rural setting and a densely populated urban setting. Second, the limited availability of **local patient-based data** is further complicated by population mobility based on a free choice of health care. The difficult vaccination demand assessment and vaccination coverage monitoring, requires a greater flexibility at the HCs and demand generation by Community Health Workers (CHWs). The granularity of the data does not correspond to the scale of the problem and structural data deficiencies, such as the Denominator error, are attributed to an inaccurate target population number. Third, **root causes** of system challenges were often diverse and distant, both in space and time, from the observed issues. A low stock level of a measles vaccine at a district hospital could be caused by higher demand due to an unforeseen campaign, recently increased local demand, a new measles vaccine introduction, a new and learning EPI responsible, or a supply problem at the manufacturer level.

The assessment at subnational level is illustrated by a typical rural and semi-urban setting, derived from cases in Rwanda and focusing on measles. Figure 6 shows clear differences between the two settings. On the one hand, the rural setting suffers from low access to the HC due to poverty (R1) and insufficient coverage of health services in the area (R2). In addition, the growing population at risk remains invisible to the surveillance and response team (B1) until an outbreak occurs (R4). On the other hand, the peri-urban setting has a high coverage of HCs, and the population visits their preferred HC. This leads to an unpredictable demand, requiring a flexible vaccine stock management at the HC (R3). Due to economically-driven population mobility and lack of patient-based immunisation data available to the HC, the local immunisation coverage rate is unknown and individual measles cases occur (R1). The surveillance & response team can intervene adequately (B1).

**Figure 6.**
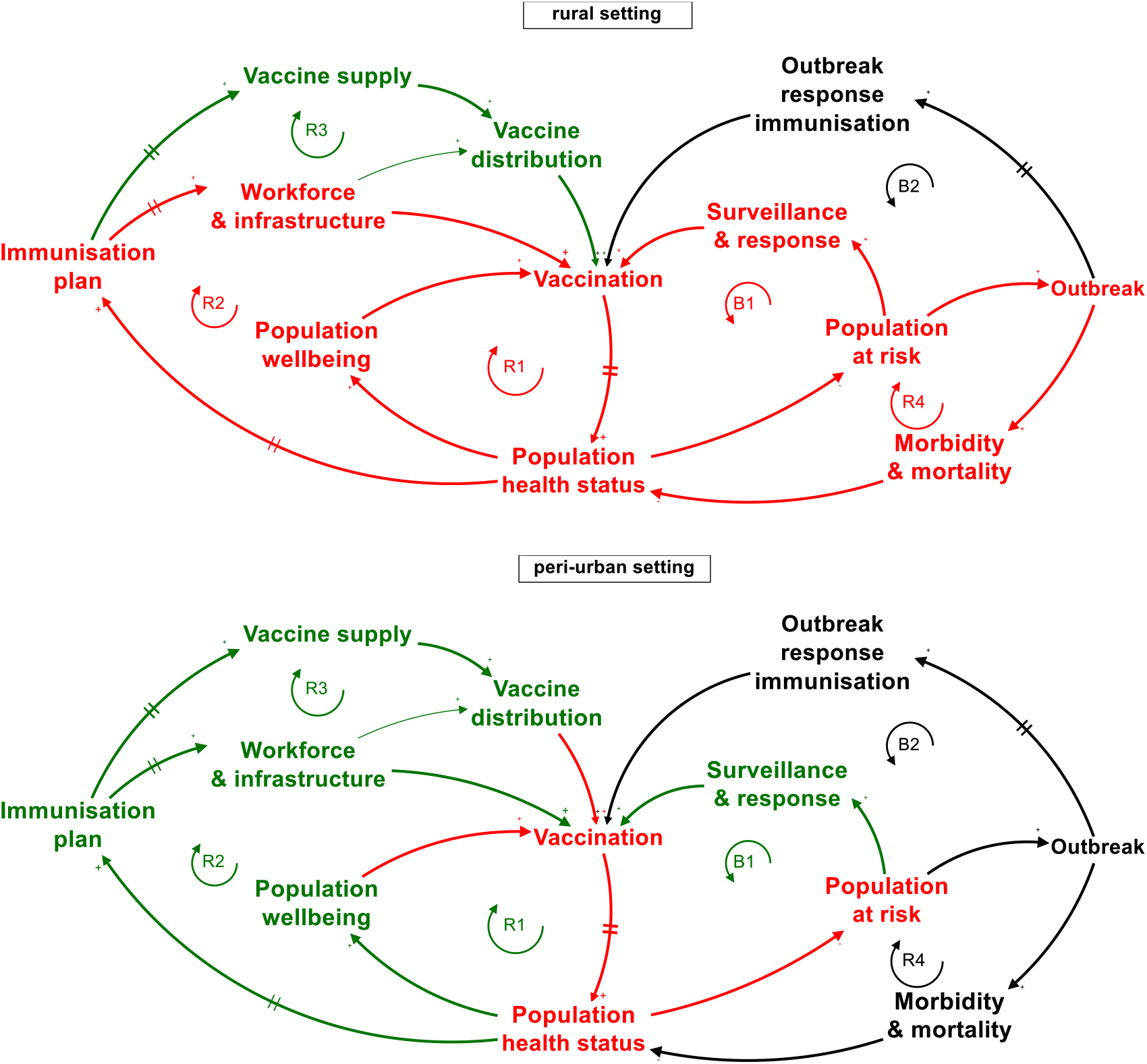
System assessment of local settings derived from cases in Rwanda: rural area (top) and peri-urban area (bottom) with respect to measles. Red loops are active with a negative impact on the IMS performance. Green loops are active with a positive impact on system performance. Black loops are not active at the moment of assessment.

Technically, the **sustainability** assessment of the IMS means investigating whether the *Planned immunisation* loops R1/R2/R3 and the *Outbreak prevention* loop B1 are capable of dominating the system till 2030 and beyond. The *Population at risk* should be kept small enough such that the surveillance & response mechanisms can avoid outbreaks. Achieving this status requires an IMS that continuously provides the right antigens needed to protect the target population from the VPDs they potentially get exposed to, in a timely and inclusive manner.

In order to assess the long term dominance of loops R1/R2/R3 and B1, the balancing loops, and factors that constrain R1/R2/R3 and B1 must be identified. This requires a structured investigation of the actual status and future evolution of the IMS system actors that keep the loops running and, equally important, the exogenous constraining factors. For example, R1 depends on (1*) health-seeking behaviour*, (2) vaccinating with the setting-specific vaccines to combat U5MR, and (3) favourable economic conditions fostering well-being. The loops R2, R3 and B1 heavily depend on the adequacy of the *Immunisation plan*, which in turn cannot be attained without *government* and *donor commitment* (Figure 7). The latter is exogenous, but with a known future evolution (GAVI graduation). It can be explicitly taken into account when designing feasible transition scenarios with increasing domestic funding.

**Figure 7.**
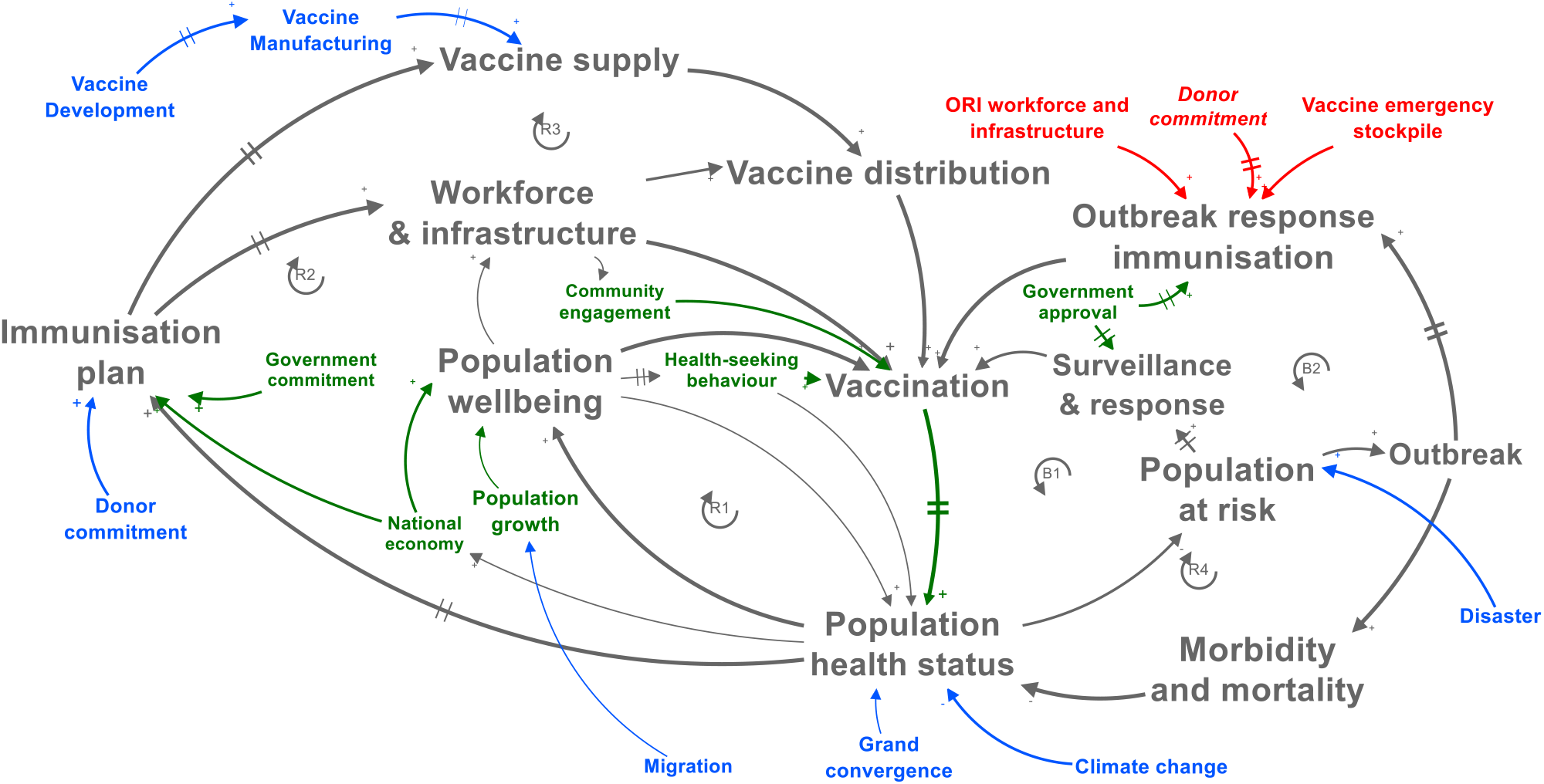
Factors determining sustainability of the IMS: endogenous factors (green) and exogenous factors (blue). Exogenous resilience factors of the IMS are red.

The evolution of *Vaccine Manufacturing* and *Vaccine Development*, the other exogenous factors driving R3, depends on the global vaccine market and the vaccine development outcomes for new pathogens.

Further exogenous factors impacting current and future demand for immunisation are (1) C*limate change*, causing reduced population health status and higher exposure to pathogens, and (2) M*igration* and its impact on population growth. Furthermore, the rise of non-communicable diseases (NCDs) and the Demographic shift in SSA will change the role of the IMS in the healthcare system. The impact of all these combined elements must be understood in order to design sustainable IMSs.

When these factors evolve disruptively, the system’s **resilience** is challenged. From the IMS diagram, the following resilience phases can be recognized: (1) <u>prevention:</u> the degree to which loops R1/R2/R3 manage to avoid the creation of a population at risk, (2) <u>mitigation:</u> the effect of loop B1 to avoid or to reduce the impact of a potential outbreak, (3) <u>response:</u> as the activation of loop B2 during an outbreak in R4, (4) <u>recovery:</u> the time and effort it takes to restart the stable mode with loops R1/R2/R3 dominating, and (5) <u>adaptation</u>: the improvements that make the IMS better equipped to cope with the next disruptive event or chronic stress.

As a consequence, the resilience of the IMS varies according to the disease type considered, the system scale, and the specific geographical setting. On top of that, the performance of loop B2 depends heavily on exogenous factors such as the timely distribution of emergency stockpiles and the availability of ORI workforce and infrastructure from humanitarian aid organizations. The rural setting in Figure 6 was clearly insufficiently resilient while the peri-urban setting showed adequately resilient behaviour.

#### Directions for system redesign

Once the root causes are identified, interventions at leverage points can be generated. Some leverage points could be identified by stakeholders, from their position on the IMS diagram during workshops and feedback meetings (activities 2 and 6 from Table 1). Potential leverage points of an IMS, classified according to the Intervention Level Framework (Malhi et al., 2009; Meadows, 1999), are listed in Table 2.

**Table 2.**
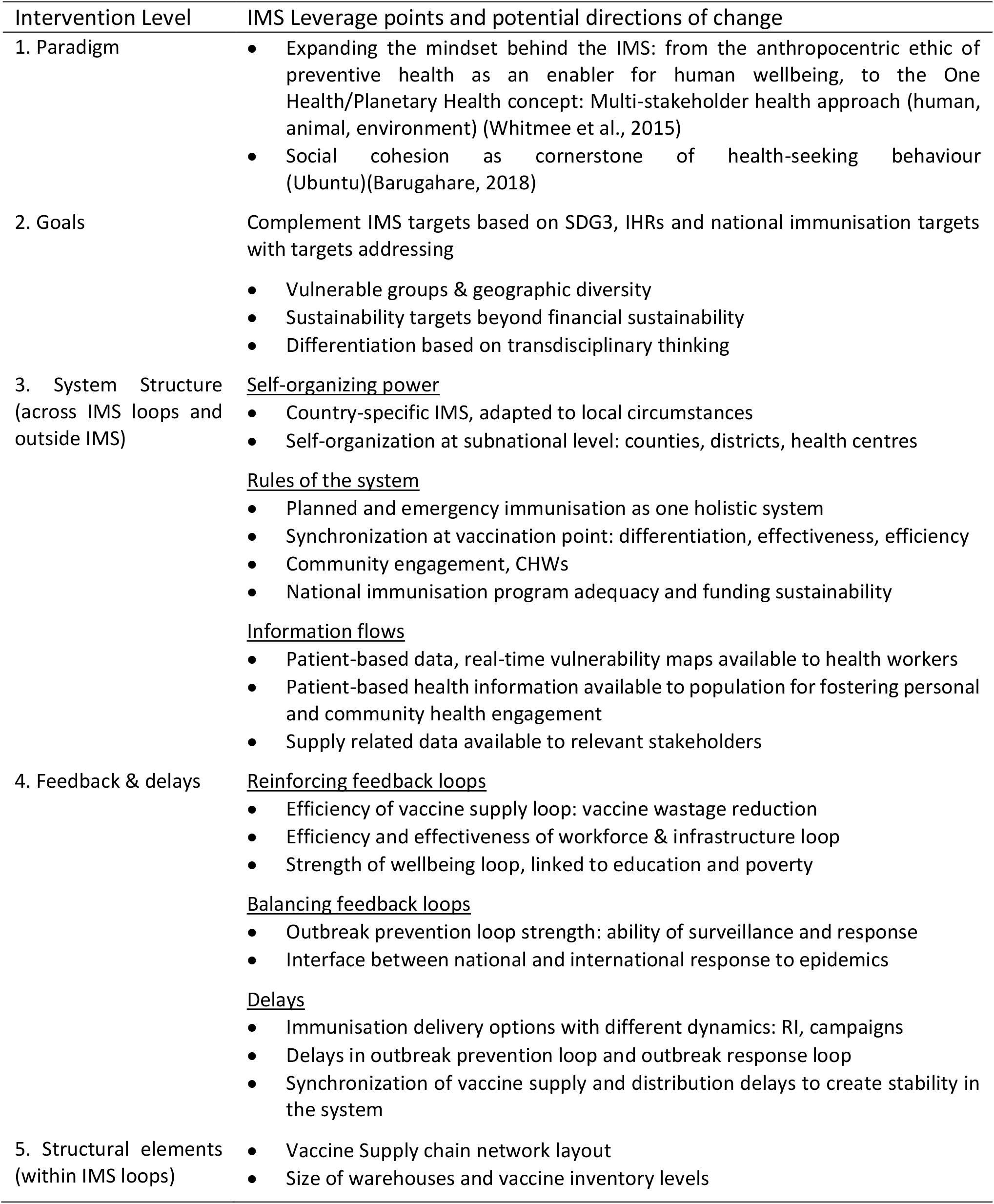
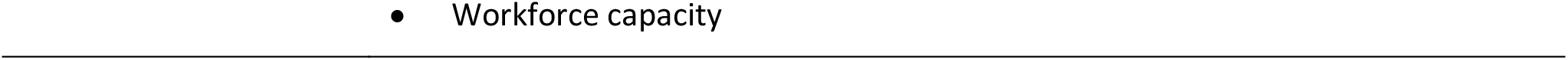
Examples of immunisation system leverage points classified according to Malhi (2009) based on Meadows (1989)

The upper categories in Table 2, such as *Paradigms* and *Goals*, are **deep** leverage points: they have a transformative and long-lasting impact but are hard to implement. **Shallow** leverage points such as structural elements are easier to implement but have a local, short term effect. The six loops that make up the IMS system represent subsystems of the IMS system. Shallow leverage points act predominantly on one subsystem, e.g. the vaccine supply loop. The more interlinkages the leverage point has with other subsystems, the higher the transformational power of the leverage point. In the rural setting in Figure 6, examples of potential interventions, at each level, are: additional staff at the nearest HC (L5), outreach vaccination at the community (L4), sustainable community-specific immunisation program, e.g. self-sustained mobile clinic (L3), IMS goals for inclusivity at the same level as national coverage (L2), and reaching a level of social cohesion in society which prevents communities/individuals to fall behind (L1).

In this way, the potential impact of interventions and the path to implement them can be modelled qualitatively. When appropriate for decision making, a numerical simulation model at (sub)national scale can be derived from the fine-tuned IMS diagrams, which were used to structure the complex system to be simulated.

A time-dependent implementation roll-out plan for a single intervention or combination of interventions constitutes an intervention scenario. Depending on the system scale, intervention scenarios appear in the health sector strategic plans, whereas the intervention implementation is captured in the cMYP, district plans or operational plans at HC level.

For the design of sustainable intervention scenarios, one can rely on (1) the knowledge that interventions at deep leverage points determine the system within which shallow leverage points operate, and (2) the concept of human-centred design to understand and model the needs and the adaptive behaviour of stakeholders.

In summary, the framework can be applied to the design of interventions and scenarios related to (1) subsystems’ structure (e.g. vaccine distribution), (2) interventions across subsystems (e.g. target setting), and (3) interventions at the boundary of the IMS (e.g. integration of immunisation with other systems).

## Discussion

Without requiring a systems background, the IMS diagram guides stakeholders along the feedback loops and paradigms that make up the complexity of an IMS. The aggregated level invites the stakeholders to use the diagram as a template, providing them with the necessary system elements to capture their mental models, generate high-leverage interventions and allow them to adapt their behaviour and decisions (Rouwette, Korzilius, Vennix, & Jacobs, 2010). In this way, the framework represents a problem structuring-step in iterative system design processes. Moreover, the framework allows to connect available findings in current literature (e.g. vaccine supply chain optimisation models) with implementation research (indicating the contextual circumstances and implementation pathway).

The proposed approach may be extended and applied to preventive health systems beyond immunisation and to infectious diseases which are not yet vaccine-preventable (e.g., nutrition, malaria, HIV, TB).

To address the problem of subnational under-immunisation, the IMS framework proposes to start from the **operational scale of the problem** instead of a larger administrative entity. As such, the heterogeneity of settings is revealed, in terms of vulnerability and health services offered. This heterogeneity may expose a need for differentiated care, or appropriate implementation models. Differentiation may lie in alternative accessibility solutions, integration with other health services, or hybrid versions of routine and campaign-based immunisation.

The notion of **sustainability** has been translated into systems behaviour: feedback loop dominance, and the evolution and impact of constraining factors. All these elements indicate whether the system is capable of evolving along with its environment to continue to fulfil its purpose in the long run. Therefore, designing for sustainability not only involves financial sustainability and increasing efficiency, but also challenges the **boundaries** of the IMS and considers interventions at deep leverage points. This creates options such as integration with preventive health systems (e.g. nutrition, family planning), interactions at the interface between immunisation and other subsystems of the health system (investment in immunisation means savings in treatment), the evolution of population growth (linked to family planning, economic development, education, migration), and the continued availability and affordability of vaccines. Because of the long term perspective, **long term phenomena** become relevant such as immune protection waning, pathogen resistance, climate-change induced disease spreading patterns, and the development of new vaccines. The design of well-coordinated sustainable response interventions requires a broader perspective related to the position of humans in the ecosystem. This broadened perspective was not addressed in this study. The integration of family planning and immunisation in one single model is a future research direction by which we want to understand the levels at which sustainable scenarios for a population health status can be obtained and if these levels are consistent with the findings of an existing socio-economic planning model such as the T21 model (Pedercini, Kleemann, Dlamini, Dlamini, & Kopainsky, 2019).

Similarly to sustainability, system **resilience** was analysed through systems behaviour. However, the time span of dynamic behaviour linked to disruptive events and resilience is much shorter (weeks or months) compared to evolutions linked to sustainability (years or decades). When the IMS is pushed to its limits, its response (loops B1 and B2) is impacted by contextual factors and effective human decision making. To fully embrace resilience in the IMS design, the transition of loop dominance from outbreak response to outbreak prevention to planned immunisation for emerging diseases should be investigated. Furthermore, the **interplay between the planned immunisation and the emergency immunisation** opens up new ways to combine their design options to jointly achieve better immunisation performance. This has not been exploited to its full potential up to now. The areas include for instance innovative types of campaigns, active engagement of target population(s) in communities, temporary settlements and mobile populations, and integration of health-promoting services.

As each **country** has its own history and **unique** political, social, economic and environmental background, the country-specific IMS has evolved and adapted to disruptive events. Many learnings, valuable for future IMS design, can be found by analysing IMSs within their specific context. Examples are the organization of community health services in Rwanda, or devolution of health care in Kenya. Additionally, this opens the door for cross-border collaboration around shared health threats, such as expanding vector habitat or migration of infected people.

This research provides a tool to investigate whether and how to push forward **deep leverage points**, as an answer to the call for sustainable IMS design. It enables to go beyond the redesign within one subsystem, and to focus on their interaction and the cornerstones of immunisation, for instance: routine immunisation, emergency response, separating EPI from health care, availability of patient-based data, and community engagement.

Limitations to this research include the number of countries and settings that served as the basis for the CLDs, and the selection of system’s concepts illustrated in this paper. In addition, the discussion of the role and integration of immunisation in the broader preventive health systems contributing to universal health coverage, and the broader sustainability dimensions of the IMS were not further explored in this paper. All of these will be included in further research.

## Conclusion

This research intended to provide a systems-inspired framework to support the design of sustainable and resilient IMSs for LMICs. Resulting from IMS stakeholder engagements, an aggregated IMS diagram was developed and applied for the assessment of immunisation performance, and for the identification of current and future challenges together with their root causes. Finally, system leverage points for IMSs were introduced to guide design directions for intervention scenarios leading to sustainable and resilient IMSs for LMICs.

The study delivered the following insights:

- The stakeholder-supported IMS diagram successfully uncovers the relation between IMS interventions and health outcomes, both on national and sub-national levels.
- In order to redesign more efficient, resilient and sustainable IMS, two paradigms, planned and emergency immunisation, should be considered as part of one IMS.
- The synchronization of three flows (child, nurse, vaccine), represented by three feedback loops, is key to IMS performance.
- The concepts of system sustainability and resilience are translated into tangible system actors and behaviour.
- A structured method was proposed to derive intervention scenarios from the conceptual IMS diagram. Consequently, the framework can be applied as a problem structuring tool as part of a system design process.

## Data Availability

The datasets generated during and/or analysed during the current study are available from the corresponding author on reasonable request.

## Acknowledgements

This work was supported by the GlaxoSmithKline Research Chair on Re-Design of Healthcare Supply Chains in Developing Countries to increase Access-to-Medicines. K. De Boeck is funded by a PhD fellowship from the Research Foundation – Flanders. The funders had no role in the design and conduct of the study. We like to thank the EPI teams in Rwanda and Kenya and all participants to workshops, interviews and field visits.

## Annex 1 Description of the six feedback loops

### (R1) The Population Wellbeing loop (VAC – PHS – PWE – VAC)

*Population Health Status* is positively impacted by the number of fully immunised children by means of *Vaccination*. The strength of this causal effect is determined by vaccine efficacy, immunological response, vaccination coverage at the community scale, and exposure to the pathogen. The effect is limited to the extent that vaccine-preventable diseases represent only part of the morbidity and mortality observed. Increased childhood vaccination is the consequence of more surviving children in the birth cohort and net immigration. Both effects are linked to population wellbeing. Access to immunisation is, in many LMIC settings, linked to the distance to the health centre, road accessibility, the cost of transportation, the education level of the caregiver, and the rank of the child in the family (Kawakatsu & Honda, 2012; Pertet et al., 2018), all elements that are positively linked to *Population Wellbeing*. Under the Reaching Every District (RED) program, outreach activities have significantly been strengthened (Bazos et al., 2015; World Health Organization, 2017a). Even with secured access, the ultimate decision for a child to get vaccinated is made by the caregiver, driven by her/his *Health-seeking behaviour*, observed as *vaccine hesitancy*. This dependency on individual human decision making is the bedrock of the need for human-centred design. The positive feedback loop is closed by the generally acknowledged relation between *Population Wellbeing* and *Population Health Status*.

### (R2) Workforce and infrastructure loop (VAC – PHS – IP – WIF – VAC)

Starting from *Population Health Status*, the same path to *Vaccination* is followed as in R1. In order for *Vaccination* to take place, a skilled vaccinator with sufficient time available to vaccinate and to manage vaccine inventories needs to be present. The vaccinator needs a data management system for vaccination coverage tracking and vaccine ordering, an accessible health centre equipped with cold chain infrastructure, transportation and equipment for outreach activities. The *Workforce and Infrastructure* dedicated to the EPI resorts under the responsibility of the Ministry of Health (MoH). The budget, strategies and interventions are planned in the national *Immunisation Plan* and, when properly done, result in better performance of the *Workforce and Infrastructure*. The restricted availability of skilled health professionals and recruitment delay adds rigidity to this loop, as well is staff retention and continued training. Similar issues are faced by the CHWs: as CHWs have access to detailed information about vaccine demand in the community, they are able to directly influence the access to immunisation. Their motivation, related to recognition, material, and financial working conditions, is, therefore, a key element (World Health Organization (WHO), 2018). Again we underscore the human-centred design aspect by the crucial role of human decision making in the hands of the immunisation workforce and the local community. The link between the *Immunisation plan* and the *Population Health Status* indicates that delivering an adequate *Immunisation plan* is more likely when the *Population Health Status* is already at a good level. With herd immunity or disease eradication already achieved, it is easier to establish a feasible *Immunisation plan* to further improve the *Population Health Status*. In contrast, a lower *Population Health Status* represents more considerable immunisation challenges that need to be addressed and a delay between the *Population Health Status* and the *Immunisation plan* exists as it takes time to collect and report the *Population Health Status* indicators such as U5MR and to plan strategies that act upon them.

### (R3) Vaccine supply loop (VAC – PHS – IP – VSU – VDI – VAC)

Upstream from the point of *Vaccination* in de CLD, the third element needed is the vaccine. Vaccine availability relies on the *Vaccine distribution* network performance, from the point of entry to the point of vaccination. The *Vaccine distribution* network includes the cold chain equipment and multiple transportation modes down to all levels where fixed vaccination and outreach take place.

Through the entire journey, the volume of vaccines, the vaccine quality, and required cold chain conditions are managed, guided by EPI procedures and WHO/UNICEF inventory guidelines. Product wastage due to expiry, cold chain excursions, and unused doses in multi-dose vials are monitored. Thanks to continued focus and support from the LMIC governments, GAVI and WHO/UNICEF and many implementing organizations, vaccine availability has increased while logistics cost and vaccine wastage are kept under control. The biggest challenge remains the last mile, where local infrastructural conditions play a huge role and where multiple health-systems strengthening programs have fueled innovative interventions. This is expressed by the causal relation between *Workforce & Infrastructure* and *Vaccine distribution*.

*Vaccine distribution* performance in the country depends on a reliable *Vaccine supply* to the LMIC. Since the beginning of IMSs, *Vaccine supply* has been the main concern that triggered actions of donor organizations. This concern is grounded in (1) *Vaccine manufacturing* (the limited availability of some vaccines) and (2) the LMICs’ ability to procure the vaccines. The first concern is caused by the complex vaccine production process, the high quality requirements and the long production lead time (Plotkin, Robinson, Cunningham, Iqbal, & Larsen, 2017). In addition, *Vaccine development* is costly and takes years, and so does the production facility construction and the approval process. Given the limited number of vaccine manufacturers and the constrained global vaccine production capacity, shortages occur. The availability of vaccines lies clearly beyond the control of individual SSA LMICs and global initiatives have been taken to further develop the vaccine market and to support production of vaccines in LMICs. The mechanism behind this is exogenous to the LMIC immunisation system. The mechanism behind this is exogenous to the LMIC IMS. The second concern related to the affordability of vaccines for LMICS, has been countered by GAVI’s vaccine funding mechanism. This mechanism supports eligible LMICs with vaccine co-financing and price negotiations. Therefore, an adequate *Immunisation plan* contains a solid vaccine funding plan, in line with the expected need for vaccines. The mechanism behind *Donor commitment* is exogenous to the LMIC system, but *Government commitment* is endogenous and its relation to the Immunisation Plan is part of the system to be understood and redesigned. Related to the Immunisation Plan it is part of the system redesign exercise. At this point, important indications of sustainability are the fraction of domestically financed vaccines and the existence of a feasible transition scenario upon GAVI graduation.

### (B1) Outbreak prevention loop (VAV – PHS – PAR – OPI – VAC)

*Outbreak prevention immunisation* is the last option to uplift low *Vaccination* levels to avoid outbreaks among the *Population at risk. Outbreak prevention immunisation* relies on a well-functioning infectious disease *Surveillance & response* (IDSR) mechanism that detects and reports cases of epidemic-prone diseases as soon as the disease-specific alert threshold is reached. If the threshold is surpassed, response actions are activated. Campaigns triggered by the IDSR team that need to be carried out urgently, are not foreseen in the NIP and may cause disruptions to the IMS, potentially harming RI activities. The latter needs to be minimized to improve resilience.

### (B2) Outbreak response loop (VAC – PHS – PAR – OBK – ORI – VAC)

The outbreak response loop is triggered by an *Outbreak* among the *Population at* risk that cannot be controlled by the national IMS. A first type of outbreak occurs when herd immunity is lost, as the outbreak could not be prevented by the planned immunisation loops R1/R2/R3 and *Outbreak prevention* loop B1. Depending on the disease, the outbreak size and the community setting, case management and immunisation activities are planned according to WHO epidemic preparedness and response guidelines. Coordination of *Outbreak response immunisation* lies with the country-level EPI staff, supported by UNICEF and NGOs if needed, or at the international level in case of a pandemic.

A second type of outbreak occurs when immunisation coverage levels were sufficiently high and the outbreak was thus not expected. The root cause can lie elsewhere: (1) the adaptive behaviour of the pathogen, (2) unobserved weaknesses in the IMS such as coverage inequity or (3) the presence of high-risk groups or undetected cases in the population. Each of these root causes urge to investigate their root causes and to improve the IMS, the vaccines, or the health system responsible for case management. These are opportunities to adapt the system to build resilience.

A third type of outbreak occurs as part of a larger public health emergency following a disaster or humanitarian crisis, for example a cholera outbreak after a flooding. The planned IMS system has not failed in its preventive performance, but the occurrence and impact of this type of outbreaks could have been smaller if the health system had been more resilient. The antigens needed in this case are usually not included in the RI schedule (e.g. oral cholera vaccine) and need to be supplied as soon as possible. The time between the detection and the first *Outbreak response immunisation* needs to be minimized, considering the acceleration of disease cases in the *Outbreak* loop. Therefore, routine procurement, production and shipping lead times do not apply. Instead, a rapid supply of vaccines during major outbreaks can be obtained through various channels: UNICEF for several childhood vaccinations; GPEI for polio; MRI for measles or rubella; the International Coordinating Group on Vaccine Provision (ICG) for meningitis, yellow fever or cholera vaccines; the Humanitarian Mechanism for pneumococcal vaccine (World Health Organization (WHO), 2017); CEPI for NTDs or delivered directly from vaccine manufacturers’ stockpiles. Furthermore, country approval and customs’ clearance constitute potential delays, human resources have to be mobilized, and cold chain equipment must be available. In order to minimize the delay before the start of an *Outbreak response immunisation* campaign, the different potential causes of delay must be tackled in parallel.

### (R4) Outbreak loop (PHS – PAR – OBK – MMT – PHS)

When *Population health status* is low and a critical group of people becomes susceptible and exposed, the *Outbreak* loop is activated and evolves according to the disease-specific epidemiological pathways. Typically, as more people get infected, contact with other people further spreads the disease. This disease transmission mechanism will continue as long as there is a group of susceptible people in the *Population at risk*. Therefore, it is paramount that the outbreak loop is counterbalanced by the *Outbreak response immunisation* loop B2 as soon as the former is activated.

